# Association of radiological severity with inflammatory biomarkers for prognostic prediction in patients with COVID-19

**DOI:** 10.1101/2023.03.14.23287265

**Authors:** Y. Zouine, M. Benzalim, S. Alj

## Abstract

**Background:** Covid – 19 caused by SARS-CoV2 has become a pandemic. It has a rapid disease progression and causes severe and fatal complications. Associating radiological severity with effective biomarkers like CRP, leucocytes, lymphocytes, D – Dimer, would be helpful in screening, categorizing patient, preventing serious complications.

**Aims and Objectives:** The aim of the study was to investigate association between levels of inflammatory biomarkers and correlate it with HRCT chest finding to identify patients at risk of fatal complications.

**Materials and Methods:** It was a retrospective monocentric observational study undertaken at Ibn Tofail hospital COVID-19 dedicated center. 177 Patients>18 year of age who were admitted from september 1, 2020 up to november 30,2020 with laboratory confirmed diagnosis of Covid – 19 were included in the study. Data was collected on demography, disease severity, laboratory measurements, radiology imaging retrospectively from records of patients. The disease severity was classified into light, mild to severe and critic based on CT Severity scoring. HRCT Chest and inflammatory biomarkers were sent in every patient at the time of admission and the outcome was recorded.

**Results:** There were 116 male patients, 61 female patients in our study. Average age of patients having severe lung involvement is 61.9years, whereas Average age of patients having non-severe lung involvement is 56.8 years and showed significant association with severity of lung involvement (p value : 0.017). Severity of lung involvement according to HRCT chest findings was greater in patients with both raised values of CRP <0.001), D – Dimer (P value 0.032) and low values of lymphocytes (P value : 0.001). Capillary oxygen SATURATION was also found to be significantly associated with radiological severity among covid-19 patients. Compared with CRP, leukocytes, lymphocytes, and D-Dimeres levels, the CT severity score had higher sensitivity, specificity, and overall accuracy in predicting severe, critical cases, and short-term mortality.

**Conclusion:** the severity of Covid – 19 disease is correlated with radiological severity andinflammatory markers thereby it will help in immediate categorization of patients into different risk groups following diagnosis, to ensure optimal resource allocation.

## INTRODUCTION

The World Health Organisation (WHO) announced that COVID-19 was a pandemic on 12 March 2020 (1). To date, 667,815,009 cases have been reported worldwide, including 6,729,542 deaths, and the daily death toll continues to rise as new variants and mutants emerge. (2).

CT was reported to have high sensitivity in patients infected by SARS-CoV-2, so it is largely used to help patient management [3] categorization of patients, helps in their clinical management, and prevention of serious complications. The degree of involvement on chest computed tomography (CT) is the most visual parameter that may reflect the severity of inflammation [4].

Biomarkers commonly evaluated to assess severity of Covid – 19 diseases are D – Dimer, CRP, IL6 and LDH.

the D-dimer level may be associated with the severity of inflammation rather than directly related to the hypercoagulable state in patients with SARS-COV-2 pneumonia. [5]

CRP in severe Covid – 19patients increased significantly at the initial stage, and is a signal of lung deterioration and progression.[6]

In this study, we examined the relationship between levels of inflammatory markers and CT severity score in patients with SARS-COV-2 pneumonia to prove whether or not there is a correlation between them, and thus to assess the role of CT versus biomarkers as a prognostic value for COVID-19 disease severity and short-term clinical outcome.

## MATERIALS AND METHODS

It was a retrospective monocentric observational study undertaken at ibn Tofail hospital COVID-19 dedicated center. 177 Patients>18 year of age who were admitted from september 1, 2020 up to november 30, 2020 with laboratory confirmed diagnosis of Covid – 19 were included in the study. Data was collected on demography, disease severity, laboratory measurements, radiology imaging retrospectively from records of patients. The disease severity was classified into light, mild to severe and critic based on CT Severity scoring. HRCT Chest and inflammatory biomarkers were sent in patients at the time of admission and the outcome was recorded

The severity of the disease was classified according to the French Society of Radiology classification which proposed a harmonisation of the estimation of the total lung extension, qualified as mild between 0-25%, moderate between 25-50%, severe between 50-75% and critical beyond 75%.

**Figure 1:**
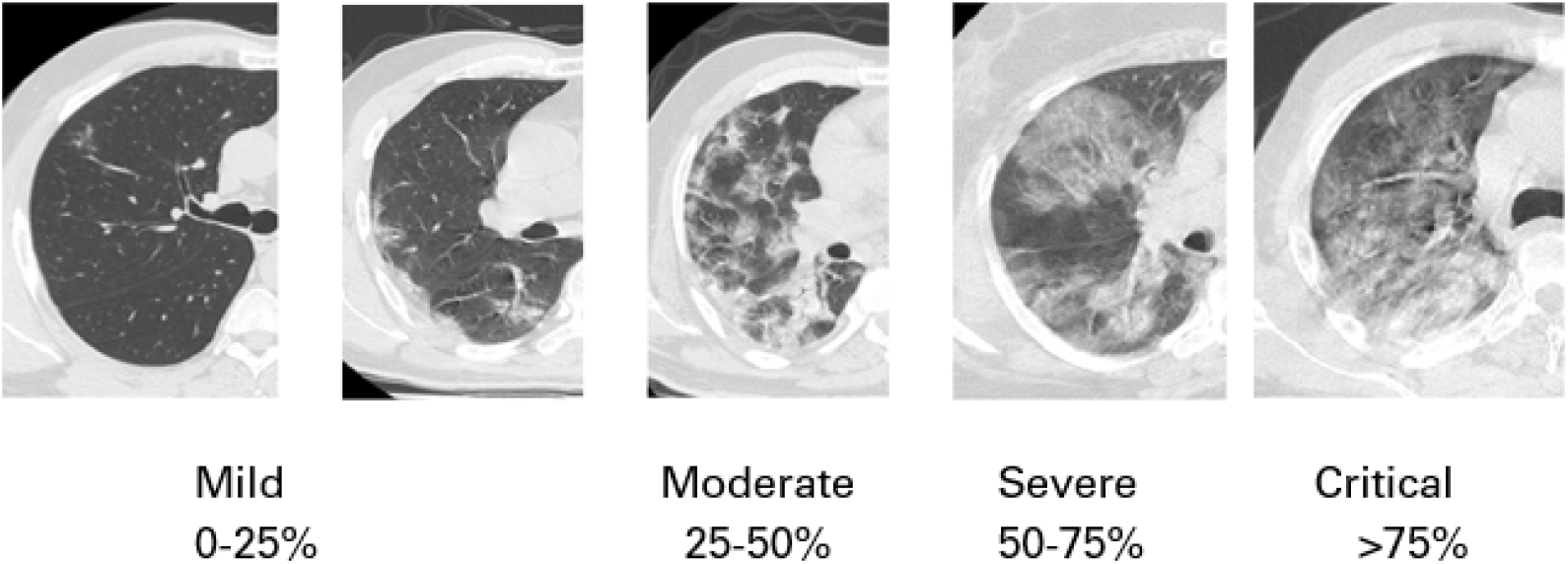
Chest CT scan demonstrating the extent of the Covid-19 lesions.

The biomarkers included in our study were the following:

**Table 1:** Reference values for inflammatory biomarkers

| BIOMARKERS | REFERENCE VALUES |
| --- | --- |
| Leukocytes | 4000-10000 elm |
| Lymphocytes | 1500-4000 elm |
| CRP | <6 mg/L |
| D-Dimères | <500 ug/L |

Statistical analysis was performed using SPSS software version 27. Descriptive analysis consisted of :

-Calculating frequencies for qualitative variables,-Using measures of central tendency (means) and dispersion (standard deviation) for quantitative variables.The t-test was used to compare means of quantitative data between 2 groups to determine whether there is a statistically significant difference between the means. For comparing means between more than 2 groups, the one-way analysis of variance (ANOVA) was used to determine whether there are any statistically significant differences between the means. Bivariate analysis was conducted to determine whether there is any relationship between 2 qualitative variables using chi-squared (χ^2^) test. The level of significance was set to 0.05 for the study.

To study the specificity and sensitivity of CT versus biomarkers, we used the ROC curve, which allowed us to specify the area under the curve of each element, called “AUC”. The AUC of the curve of the biomarker studied must be greater than or equal to 0.5 in order for it to perform well. The ROC curves also allow the detection of the best CT threshold value and biomarkers for the detection of severe cases.

## RESULTS

There were 116 male patients, 61 female patients in our study.

Average age of patients having severe to critic lung involvement is 61.9, whereas Average age of patients having light to mild lung involvement is 56.8 (on comparison of means, p value 0.017). A significant association between age and severity of lung involvement was found

We showed that there was a significant reverse relationship between CT severity score and capillary oxygen saturation (p value <0.001)

Out of 177 patients that we have included in our study, 28 patients with severe to critical lung involvement had raised D –Dimer. (p value=0.032)

In our study, there were 146 patients with raised CRP, out of which 78 were having severe lung involvement and 68 had light to mild lung involvement. In contrast, there were 31 patients with normal CRP levels, of which 10 had severe lung involvement and 21had non-severe lung involvement. There was significant association between levels of CRP and severity of lung involvement (p value p<0.001)

There were 136 patients with lymphopenia, out of which 71 were having severe lung involvement and 65 had non-severe lung involvement. In contrast, there were 41 patients with normal lymphocytes levels, of which 9 had severe lung involvement and 32 had non-severe lung involvement. There is significant association between levels of lymphopenia and severity of lung involvement (p value =0.001)

Out of 177 patients that we have included in our study, 49 patients had isolated raised creatinine. It showed weak association with severe lung involvement (p=0.207).

In our study, the calculated AUCs were as follows: age 0.663, saturation 0.797, leukocytes 0.572, lymphocytes 0.682, DD 0.651, CRP 0.746. Biomarkers with AUC< or equal to 0.5 were excluded from the next step (Creatinine).

ROC curve analysis showed that the area under the curve (AUC) was significantly elevated using CT severity score cut-off ≥ 50%, CRP cut-off ≥ 95 mg/L, leukocyte cut-off ≥ 11210 elm/mm^3^ and DD cut-off ≥ 700 ug/L for severe COVID-19 cases, with a sensitivity, specificity of 85% 58%, compared with 75% 55% for CRP, 85% 55%, compared with 62% 55% for leukocytes and 80% 48% compared with 70% 40% for DD respectively.

## DISCUSSION

Lung computed tomography (CT) scan is an important method in the diagnosis of pulmonary abnormalities, which also has a very valuable role in screening the patients suspected of infections as well as diagnosis and clinical classifications of these patients.[7]

The most common patterns of this infection are bilateral and multilobar ground-glass opacities.[8,9] It has also been shown that the peripheral and lower lobes of the lung are mostly affected. Further studies also showed other radiological patterns, such as crazy paving pattern, airway change, and reverse halo sign for patients infected with COVID-19.[10,11]

Some studies have also emphasized the relationship between CT scan findings and the clinical condition of patients explaining the possible role of lung CT scan in determining the severity and spread of the disease[12]

The aim of our study is to assess the severity of Covid – 19 disease based on its correlation with level of biomarker and radiological severity

CRP is a non-specific acute-phase protein induced by IL-6 in liver and sensitive biomarker of inflammation, infection, and tissue damage.18 Studies showed that it increased significantly in severe Covid – 19 patients at the initial stage, which is a signal of lung deterioration and disease progression as in Prakhar and al study that demonstrated a significant association between levels of CRP and severity of lung involvement (p=0.0346, RR of 2.02, Odds Ratio of 2.37).11 Our study agrees and confirms that CRP levels can be an indicator for severe disease and progressive inflammation.[13,14]

Several studies also demonstrated that a higher level of D-dimer was associated with in-hospital mortality[15,16]

The most suggested mechanism was that the hypercoagulable state, which could be reflected by an elevated D-dimer level, might lead to thrombotic events, resulting in poor outcomes. However, the coagulopathy was thought to result from local and systemic inflammation caused by the coronavirus. Also, D-dimer is known as a biomarker of inflammation [17]. Therefore, LAN WANG and al proposed the hypothesis that the D-dimer level may be associated with theseverity of inflammation rather than directly related to the hypercoagulable state in patients with SARS-COV-2pneumonia.And showed that Patients with D-dimer level > 0.7 mg/L had significantly higher CT score (12.0 [P < 0.001) and higher incidence of reticulation and/or traction bronchiectasis on chest CT images (83.3% vs. 46.2%, P = 0.002) than patients with D-dimer level ≤ 0.7 mg/L. The natural logarithm of the D-dimer level was significantly associated with the CT score (rS = 0.586, P < 0.001) [5]

Lymphopenia was reported in varies types of virus-infected diseases, such as SARS [18, 19, 20], MERS [21, 22] and respiratory syncytial virus [23]. A previous study reported that lymphopenia in SARS may be due to enhanced vascular sequestration associated with increased soluble vascular cell adhesion molecule-1 levels [24]

In our study lymphopenia was a good predictor of severe lung involvement wich agrees with the study of Jiheng Liu and al that showed the correlation of Lymphopenia with severity grades of pneumonia (P<0.001). Lymphopenia was associated with a prolonged duration of hospitalization (17.0 days vs. 14.0 days, P = 0.002).[25]

## CONCLUSION

Older age (>61 years), elevated D-dimer, CRP and low lymphocyte count are significantly correlated with severe lung injury and are risk factors for poor prognosis in patients with COVID-19. CT severity scoring has high accuracy and significant predictive power compared to other biomarkers. In particular, it has higher sensitivity, specificity and accuracy than CRP, WBC, lymphocytes and D-dimer. Our study revealed that CT scoring is a powerful means to determine the risk of death from COVID-19 disease in patients and to triage the need for hospitalisation. It is also useful for prioritising medical resources to make better decisions for the best clinical outcomes.

## FIGURES AND TABLES

**Table 2:** Comparison of capillary blood saturation between patients who have light, mild, severe and critical lung involvement.

|  | n | Mean Sao2 | Standard deviation | F | p |
| --- | --- | --- | --- | --- | --- |
| Light | 39 | 95.0513 | 3.23589 | 29.436 | <0.001 |
| Mild | 46 | 92.2609 | 4.28659 |  |  |
| Severe | 62 | 88.1452 | 6.50560 |  |  |
| Critical | 26 | 82.9615 | 7.53913 |  |  |

**Table 3:**
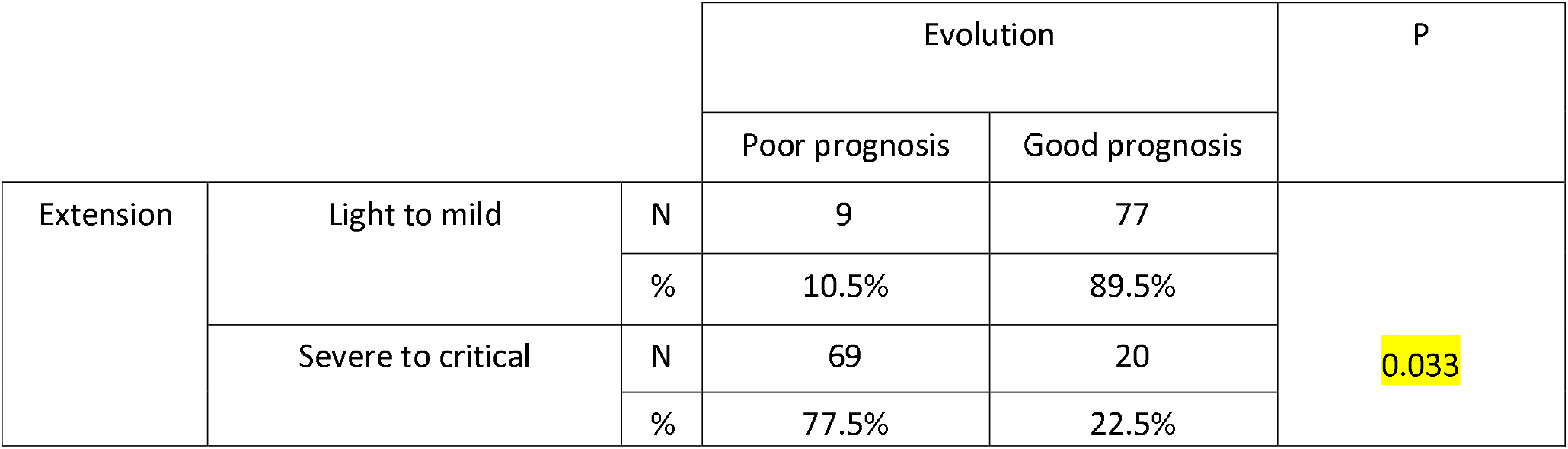
Association of prognosis and lung involvement.

**Table 4:** Comparison of Age, capillary blood saturation and biological findings between patients who have light to mild, and severe to critical lung involvement.

|  | Extension | N | Mean | Standard deviation | p |
| --- | --- | --- | --- | --- | --- |
| Age | Light to mild | 86 | 56.85 | 16.123 | 0.017 |
|  | Severe to critical | 89 | 61.91 | 11.163 |  |
| Sao2 | Light to mild | 85 | 93.54 | 4.067 | p<0.001 |
|  | Severe to critical | 88 | 86.61 | 7.188 |  |
| Leucocytes | Light to mild | 67 | 9244.18 | 3585.765 | 0.13 |
|  | Severe to critical | 77 | 11670.13 | 12610.690 |  |
| CRP | Light to mild | 68 | 87.46 | 72.483 | p<0.001 |
|  | Severe to critical | 78 | 161.08 | 120.679 |  |
| Lymphocytes | Light to mild | 65 | 1351.8480 | 669.07303 | 0.001 |
|  | Severe to critical | 71 | 998.6338 | 589.55429 |  |
| CREAT | Light to mild | 62 | 8.44 | 3.159 | 0.216 |
|  | Severe to critical | 66 | 9.81 | 8.133 |  |
| TP | Light to mild | 18 | 28.389 | 3.5721 | 0.88 |
|  | Severe to critical | 39 | 28.274 | 2.0953 |  |
| DD | Light to mild | 34 | 1479.368 | 1386.6173 | 0.032 |
|  | Severe to critical | 28 | 5757.036 | 9932.4269 |  |

**Table 5:** Comparison of Age, and biological findings with capillary blood saturation.

|  | Sao2 | N | Mean | Standard deviation | p |
| --- | --- | --- | --- | --- | --- |
| Age | <90 | 63 | 62.94 | 11.961 | 0.012 |
|  | >=90 | 112 | 57.38 | 14.830 |  |
| Leucocytes | <90 | 49 | 10749.80 | 5095.902 | 0.843 |
|  | >=90 | 94 | 10412.87 | 11313.677 |  |
| CRP | <90 | 52 | 179.65 | 125.432 | <0.001 |
|  | >=90 | 93 | 97.96 | 83.360 |  |
| Lymphocytes | <90 | 44 | 980.0682 | 651.04801 | 0.23 |
|  | >=90 | 90 | 1253.6680 | 641.13675 |  |
| CREAT | <90 | 43 | 9.52 | 5.159 | 0.706 |
|  | >=90 | 83 | 9.07 | 6.799 |  |
| TP | <90 | 17 | 28.453 | 2.2584 | 0.792 |
|  | >=90 | 40 | 28.250 | 2.7819 |  |
| DD | <90 | 26 | 6136.923 | 10217.6589 | 0.029 |
|  | >=90 | 35 | 1465.300 | 1379.6588 |  |

**Table 6:** Association of prognosis and capillary blood saturation.

|  | Evolution | n | Mean | Standard deviation | *p* |
| --- | --- | --- | --- | --- | --- |
| Sao2 | Good prognosis | 31 | 83.9355 | 8.38226 | <0.001 |
|  | Bad prognosis | 144 | 91.2014 | 5.77138 |  |

**Figure 2:**
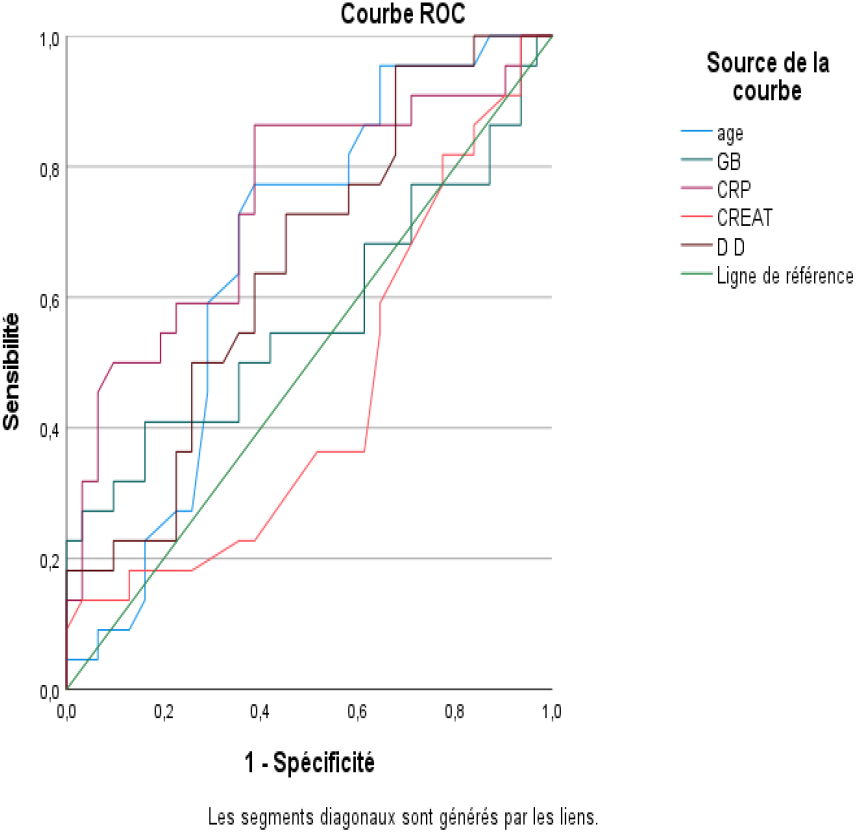
ROC curve.

**Table 7:**
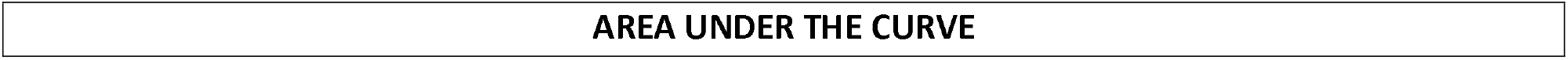
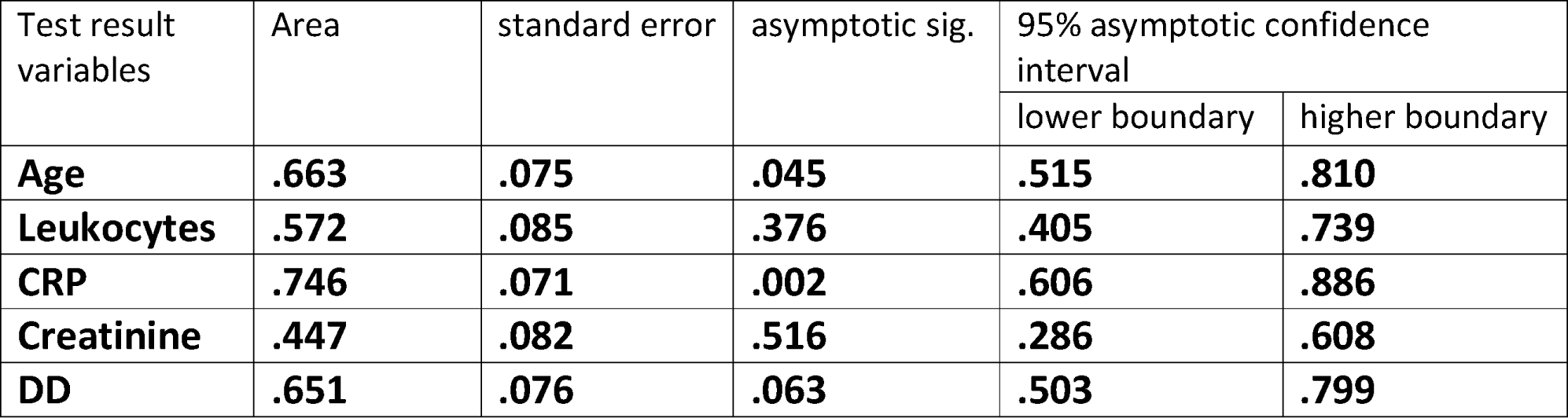
Areas under the curve of the variables.

| Test result variables | Area | standard error | asymptotic sig. | 95% asymptotic confidence interval |  |
| --- | --- | --- | --- | --- | --- |
|  |  |  |  | lower boundary | higher boundary |
| **Age** | **.663** | **.075** | **.045** | **.515** | **.810** |
| **Leukocytes** | **.572** | **.085** | **.376** | **.405** | **.739** |
| **CRP** | **.746** | **.071** | **.002** | **.606** | **.886** |
| **Creatinine** | **.447** | **.082** | **.516** | **.286** | **.608** |
| **DD** | **.651** | **.076** | **.063** | **.503** | **.799** |

**Figure 3:**
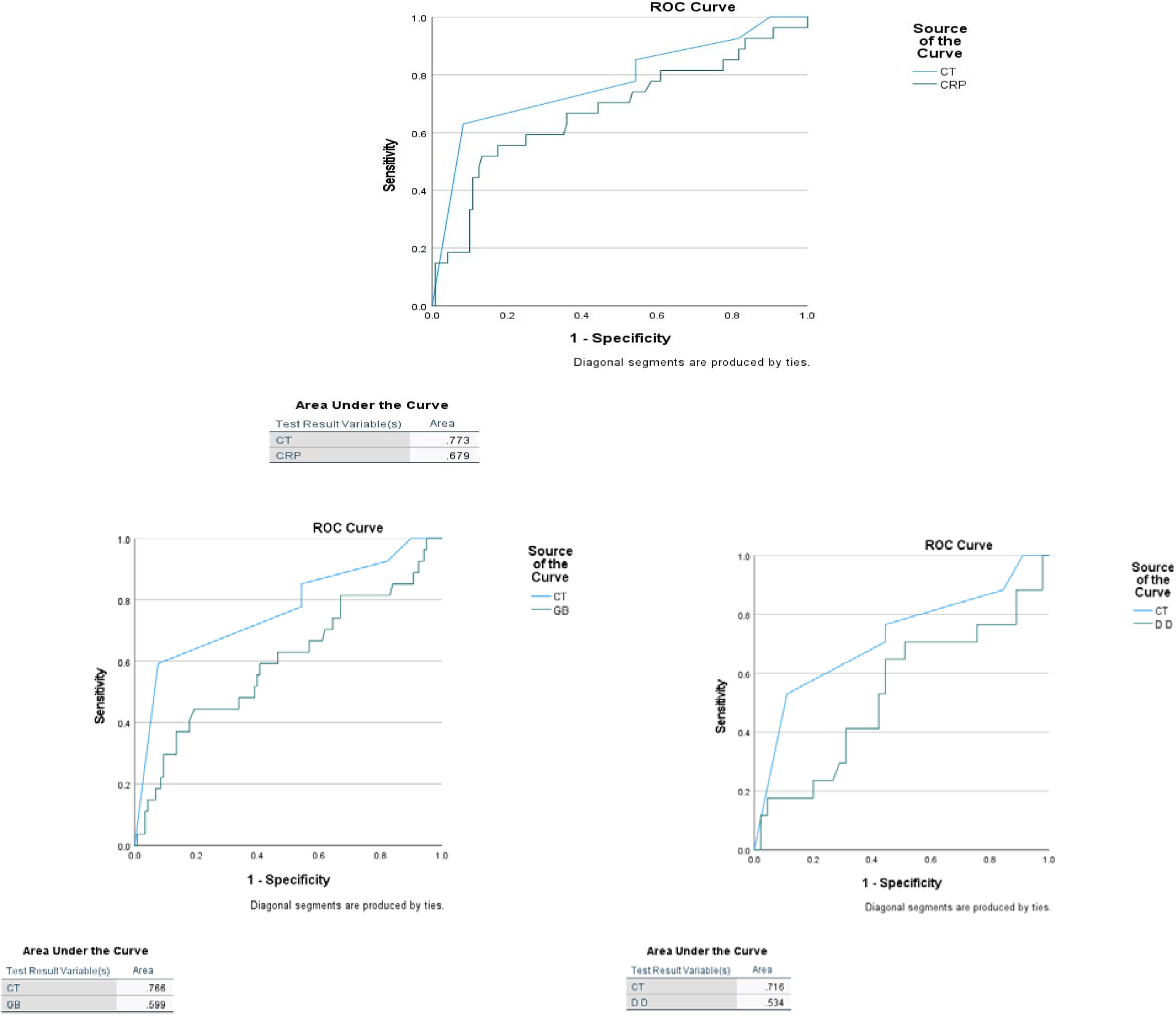
ROC curve using CT extension and CRP, leukocytes and DD levels.

## Data Availability

All data produced in the present study are available upon reasonable request to the authors

## Notes

### Competing Interest Statement

The authors have declared no competing interest.

### Funding Statement

This study did not receive any funding

### Author Declarations

the ethics committee at the mohamed 6 university hospital

